# Tune Out: A randomised controlled trial to investigate the impact of an online program on tinnitus severity, handicap, and psychological symptoms in adults with tinnitus

**DOI:** 10.64898/2026.07.05.26357341

**Authors:** Emma Laird, Daniel Gosbell, Adriana L. Dall’Est, Alicja Malicka

## Abstract

**Objective:** To evaluate the efficacy, engagement, and usability of Tune Out, an unguided, self-paced online tinnitus management program, for reducing tinnitus severity in adults with tinnitus.

**Design:** A two-arm, parallel-group randomised controlled trial was conducted with Australian adults reporting diagnosed or self-reported tinnitus. Participants were randomised to immediate access to Tune Out or a waitlist control group. Outcomes were assessed at baseline, 6 weeks, and 12 weeks. The primary outcome was tinnitus severity measured using the Tinnitus Functional Index (TFI). Secondary outcomes included tinnitus handicap, psychological symptoms, program engagement, self-efficacy, and usability.

**Results:** Eighty-eight participants were randomised: 43 to the intervention group and 45 to the waitlist control group. The primary outcome analysis included 63 participants at 12 weeks. A significant Group × Time interaction was observed for TFI total score, indicating greater reductions in tinnitus severity over time in the intervention group compared with waitlist control, *F*(2, 102.57) = 5.95, *p* = .004, partial η² = .104. Significant effects were also observed for tinnitus handicap, *F*(2, 106.76) = 4.12, *p* = .019, partial η² = .072. Effects on psychological symptoms were less consistent, although anxiety showed a significant Group × Time interaction, *F*(2, 116.85) = 3.63, *p* = .030, partial η² = .059. At 12 weeks, 23.1% of intervention participants achieved a clinically meaningful reduction in tinnitus severity compared with 5.4% of controls. Program use was highly variable, with a median use of 1.10 hours, and 25.6% of intervention participants recording no use. Usability ratings were favourable among respondents, with a mean System Usability Scale score of 73.13.

**Conclusions:** Tune Out demonstrated preliminary efficacy for reducing tinnitus severity and tinnitus handicap compared with waitlist control. Effects on broader psychological symptoms were less consistent. Although usability was rated positively, low and variable engagement highlights the need for strategies to support uptake and sustained use in unguided digital tinnitus interventions.

## INTRODUCTION

Tinnitus is the phantom perception of sound in the absence of external auditory stimuli, typically described as a tonal ringing, buzzing or rushing noise (Baguley et al., 2013). Whilst often transient, it can develop into a chronic condition which is notoriously difficult to manage given no consistently effective cure to eliminate or reduce its severity. A meta-analysis estimated the prevalence of chronic tinnitus at 9.8% (95% CI, 4.7% – 19.3%) of the adult population (Jarach et al., 2022). Whilst many people will successfully habituate to chronic tinnitus, there is a subset of approximately 2.3% (95% CI, 1.7% – 3.1%) of people living with severe tinnitus, causing significant distress and impacts on daily life (Andersson, 2002; Jarach et al., 2022). People with severe tinnitus have significantly higher rates of anxiety, depression, and insomnia, which can exacerbate symptoms, making it a distressing condition placing a considerable burden on patients (Bhatt et al., 2017; Jiang et al., 2025; Langguth et al., 2013; Pavaci et al., 2019).

Tinnitus management varies widely across countries and services, with inconsistent guidelines and limited access to evidence-based care (Cima et al., 2020; Langguth et al., 2023). Common approaches include informational counselling, wax management, sound therapy, hearing devices for those with co-occurring hearing loss, and medical referral (Bhatt et al., 2016); however, evidence for many interventions remains limited by heterogeneity in tinnitus presentation, measurement challenges, placebo and nocebo effects, and methodological limitations in the research (Hoare et al., 2011).

Increasingly, tinnitus severity is understood not only in terms of the acoustic perception itself, but also the individual’s psychological response to the sound, often referred to as tinnitus distress (Bruggemann et al., 2016). This can include heightened awareness, stress responses, negative interpretations of the sound, and associated anxiety or low mood, which contribute substantially to functional impact (Andersson & Westin, 2008).

Psychological interventions therefore aim to reduce tinnitus distress by modifying the person’s response to tinnitus rather than eliminating the sound. Cognitive Behavioural Therapy (CBT) has the strongest evidence for reducing tinnitus-related distress and associated symptoms and is commonly recommended in contemporary reviews and guidelines (Cima et al., 2014; Henry et al., 2022; Langguth et al., 2023). Related approaches, such as Tinnitus Retraining Therapy (TRT) and Acceptance and Commitment Therapy (ACT), also target habituation, coping, and acceptance (Phillips & McFerran, 2010; Ungar et al., 2023; Westin et al., 2011). Despite this evidence, access to tinnitus-specific CBT remains limited. Psychologists and other mental health professionals may have little tinnitus-specific training, while audiologists, who are often the first point of contact for tinnitus care, may have limited training in CBT or mental health support (Henry et al., 2022). In Australia, access may be further constrained by funding models that provide limited coverage for psychological tinnitus care, leaving many patients to self-fund treatment (Patel et al., 2022). These barriers highlight the need for accessible, scalable models of psychological tinnitus support, such as CBT delivered online (Weise et al., 2016).

Internet-delivered CBT (iCBT) offers a non-invasive and accessible alternative to face-to-face CBT, particularly for people who are geographically isolated, have limited access to trained clinicians, or face financial barriers to care. Current evidence suggests that iCBT can reduce tinnitus distress, with meta-analytic findings showing large effects for tinnitus distress and smaller but significant improvements in tinnitus handicap, depression, anxiety, and insomnia in both the short and longer term (Sattel et al., 2025). Economic modelling also suggests that iCBT is likely to be cost-effective compared with minimal care, such as a single GP appointment, and is second only to group-based CBT in cost-effectiveness (Patel et al., 2022).

A key limitation of iCBT is adherence, with some studies reporting high attrition rates that limit interpretation of findings (Abbott et al., 2009; Beukes et al., 2021). Unguided iCBT may further improve accessibility because it requires little or no clinician input and avoids scheduled guidance sessions. Evidence indicates that unguided iCBT can reduce tinnitus severity and distress (Demoen et al., 2023), and one study reported lower attrition in unguided iCBT than guided iCBT, with lack of time identified as a common barrier to participation (Rheker et al., 2015). Internet delivery may also be suitable for other psychological tinnitus interventions. For example, internet-delivered ACT has been found to be similarly effective to iCBT in reducing tinnitus severity, suggesting that acceptance-based approaches may provide another accessible pathway for psychological tinnitus care (Hesser et al., 2012). However, the evidence base for unguided iCBT and related therapies remain small, and further research is needed to establish its effectiveness more conclusively (Beukes et al., 2019; Demoen et al., 2023).

Given the potential challenges of adherence in unguided iCBT for tinnitus, it is important to consider factors that may influence participants’ capacity to engage with self-guided management. Self-efficacy refers to an individual’s belief in their ability to perform behaviours required to achieve a desired outcome (Bandura, 1977), and in rehabilitation contexts it may support motivation, persistence, and engagement with behaviours needed to achieve rehabilitation goals (Bandura, 1977; Gangwani et al., 2022). In internet-delivered therapy, self-efficacy has been conceptualised as confidence in mastering obstacles during treatment, and may be relevant to both symptom reduction and attrition (Schønning & Nordgreen, 2021). However, evidence regarding the role of self-efficacy in tinnitus-specific internet interventions remains limited, particularly in relation to unguided programs.

This study extends the existing iCBT literature by evaluating Tune Out, an unguided, self-paced online tinnitus management program. The primary aim was to determine whether Tune Out reduced tinnitus severity compared with a waitlist control group. Secondary aims were to examine the impact of Tune Out on tinnitus handicap and psychological symptoms; explore whether program engagement was associated with self-efficacy or changes in tinnitus and psychological outcomes; and evaluate the usability of the program. It was hypothesised that participants allocated to Tune Out would demonstrate greater improvements in tinnitus outcomes than participants allocated to the waitlist control group.

## METHODS

### Study design

This study was a two-arm, superiority, parallel group (one to one allocation ratio) randomised-controlled trial evaluating the effectiveness of the Tune Out program. The reporting of the trial was guided by the CONSORT 2025 statement for randomised trials, and a completed CONSORT 2025 checklist is provided as Supplementary File 1. This study was registered with the Australian New Zealand Clinical Trials Registry (ACTRN12625000424404P). Ethical approval was obtained from the La Trobe University Human Research Ethics Committee (HEC25138). All participants provided informed consent before taking part, and all study procedures were conducted in accordance with approved ethical protocols.

### Participants

Participants were recruited between June and October 2025 via convenience and snowball sampling methods from a population of Australian adults who reported lived experience of tinnitus. Advertising and recruitment occurred via social media advertisements in tinnitus community groups and via the Tinnitus Awareness electronic mailing list of the Australian charity, Soundfair. The study was completed in a solely online setting.

Eligibility criteria required participants to be aged 18 years or older, currently reside in Australia, have internet access, and report current diagnosed or self-reported tinnitus. Participants were also required to have previously consulted an audiologist or medical practitioner regarding their tinnitus. Individuals were excluded if physical, linguistic, or intellectual factors prevented them from providing informed consent. These criteria were informed by the terms and conditions of the publicly available Tune Out program (Tinnitus Services Pty Ltd., 2024).

An a priori power analysis was performed in G*Power 3.1.9.7 (Faul et al., 2007) using the primary treatment outcome of tinnitus severity as measured by the Tinnitus Functional Index (TFI), which suggested a total sample size of N=86 (approx. N=43 in each group) was required to detect a medium effect size (Cohen’s f = 0.25) at an alpha of 0.05 and power of 0.8 using repeated-measures, between-factors ANOVA.

### Procedure

Prospective participants accessed the screening and enrolment survey by following a link provided in study advertisements. The REDCap online survey collected basic screening information and included the plain language statement and consent form. Participants who did not meet the eligibility criteria, or who did not wish to proceed, were notified at the point of survey submission via redirection to a non-eligibility webpage. Eligible participants were automatically redirected to the baseline questionnaire, also hosted in REDCap. Baseline measures included the Tinnitus Handicap Inventory (THI), Tinnitus Functional Index (TFI), Depression Anxiety and Stress Scale–21 items (DASS-21) and General Self-Efficacy Scale (GSE). These baseline data were considered Timepoint 0 (T0).

Following completion of the baseline questionnaire, participants were randomised to the intervention or waitlist control group using simple randomisation. Randomisation was automated within REDCap using randomisation module and an uploaded allocation table generated by Sealed Envelope Ltd. (2024). The trial used a single-blind design, whereby group allocation was concealed from the researchers but not from participants, as participants were necessarily aware of whether they had received immediate access to the program. Following randomisation, participants received an automated email advising them of their allocation and access arrangements.

Participants allocated to the intervention group received an immediate link and access code for the Tune Out program. Participants in the intervention group were encouraged to complete the program at their own pace, at a time and place of their choosing, across a three-month period. Participants allocated to the waitlist control group were informed that they would receive access to Tune Out after three months, once data collection was complete.

No minimum amount or frequency of program use was required; participants could engage with the modules and interactive activities as much or as little as they wished. At six weeks post-randomisation, Timepoint 1 (T1), and three months post-randomisation, Timepoint 2 (T2), participants in both the intervention and waitlist control groups were asked to complete the follow-up questionnaires, which included repeat administration of the TFI, THI, DASS-21 and GSE measures as well as the System Usability Scale (SUS) at T1 to assess usability of the online platform. Participant flow is presented using the CONSORT flow diagram in Figure 1.

**Figure 1.**
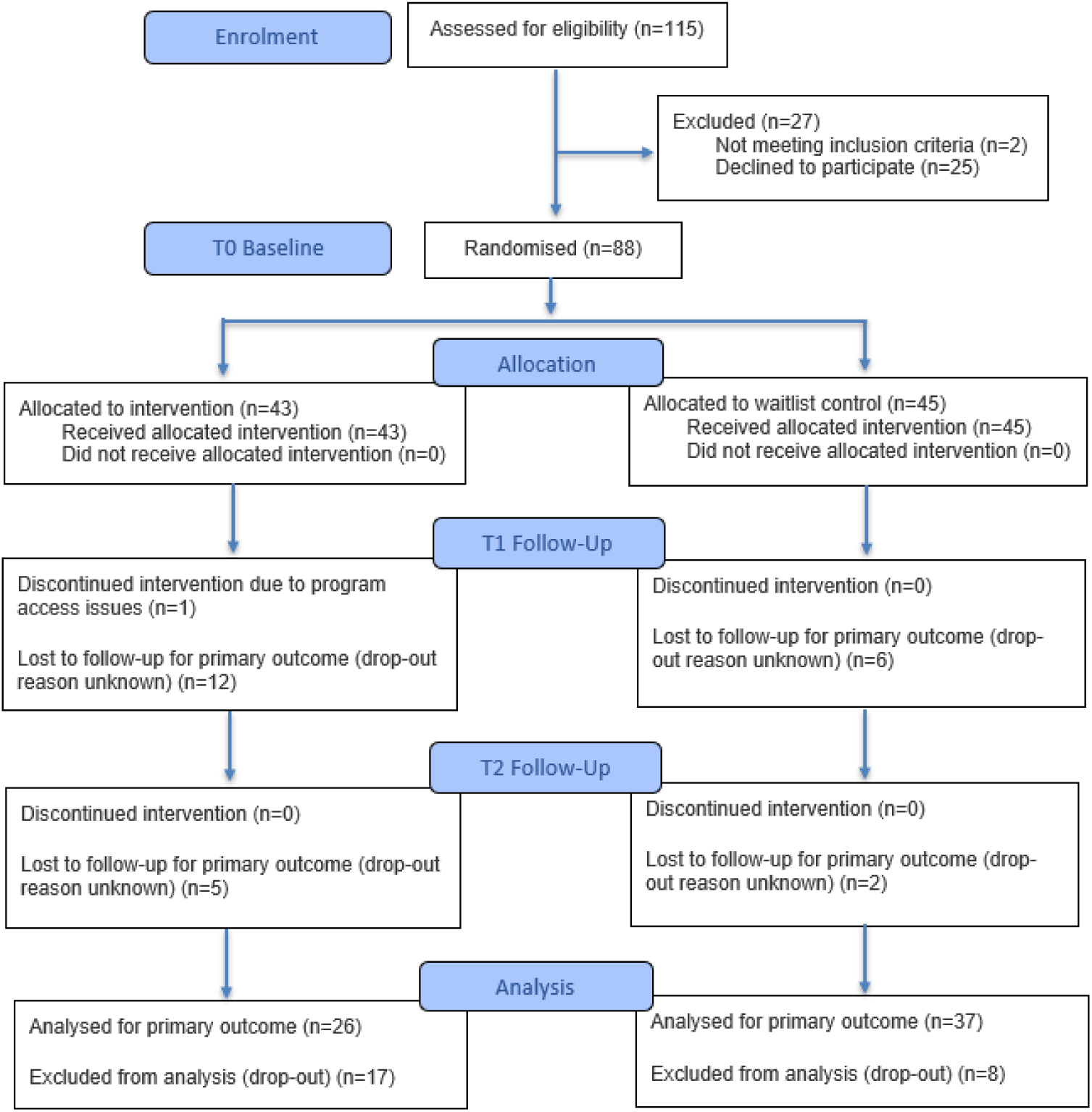
CONSORT 2025 Flow Diagram.

### Intervention

Tune Out is an unguided, self-paced online tinnitus management program consisting of 10 short modules. The program was developed psychologists with clinical experience supporting people with tinnitus. It is primarily based on CBT principles and incorporates elements of ACT. The program provides tinnitus-related education alongside practical strategies designed to help users understand, respond to, and manage tinnitus more effectively.

The 10 modules cover: tinnitus causes, impacts, and treatment options; the hearing system; understanding and breaking the cycle of problem tinnitus; CBT for tinnitus; changing thoughts and feelings in response to tinnitus; changing behaviours in response to tinnitus; reducing stress and psychological arousal; using sound to manage tinnitus; tinnitus and sleep; and ongoing tinnitus management. Interactive journal activities are included throughout the program to support reflection and application of strategies between modules.

### Measures

The Tinnitus Functional Index (TFI) (Meikle et al., 2012) was used as the primary outcome measure of tinnitus severity. The TFI is a 25-item questionnaire that produces an overall score from 0 to 100, with higher scores indicating greater tinnitus severity. It includes eight subscales: intrusiveness, sense of control, cognitive interference, sleep disturbance, auditory difficulties, relaxation, quality of life, and emotional distress. The TFI has demonstrated strong internal consistency, test–retest reliability, and responsiveness to treatment-related change (Meikle et al., 2012).

Secondary outcomes included tinnitus handicap, psychological symptoms, digital intervention usability, and program usage. The Tinnitus Handicap Inventory (THI) (Newman et al., 1996) was used to assess tinnitus-related handicap. The THI is a 25-item questionnaire that provides a total score and three subscale scores reflecting functional limitations, emotional responses, and catastrophic reactions, with higher scores indicating greater tinnitus-related handicap. The THI is widely used in tinnitus research and has demonstrated good reliability and validity (Newman et al., 1996).

The Depression Anxiety Stress Scale–21 items (DASS-21) (Lovibond & Lovibond, 1995) was used to assess psychological symptoms. The DASS-21 includes three seven-item subscales measuring depression, anxiety, and stress, with higher scores indicating greater symptom severity. The DASS-21 has demonstrated good internal consistency across clinical and non-clinical populations (Lovibond & Lovibond, 1995; Osman et al., 2012).

The General Self-Efficacy Scale (GSE) (Schwarzer & Jerusalem, 1995) was used to assess perceived self-efficacy. The GSE is a 10-item scale, with higher mean scores indicating greater perceived capacity to manage difficult or stressful situations. The scale has demonstrated good reliability and cross-cultural validity (Schwarzer & Jerusalem, 1995).

The System Usability Scale (SUS) (Brooke, 1996) was used to assess perceived usability of the Tune Out program at T1. The SUS is a 10-item measure that produces a score from 0 to 100, with higher scores indicating greater perceived usability. It is widely used to evaluate digital systems and has demonstrated robust psychometric properties across technology contexts (Brooke, 1996). Program usage data were also collected at T2 using web analytics, including number of program accesses, duration of use, and modules completed.

### Analysis

Descriptive statistics, including frequencies and percentages for categorical variables and means, standard deviations, medians and ranges for continuous variables, were summarised for participant characteristics and outcome measures. Baseline differences between the intervention and waitlist control groups, and differences between participants who completed follow-up assessments and those who withdrew or were lost to follow-up, were examined descriptively and using appropriate inferential tests. Continuous variables were compared using independent samples *t*-tests where assumptions of normality and homogeneity of variance were met. Where continuous data were non-normally distributed, Mann–Whitney *U* tests were used. Categorical variables were compared using Pearson’s chi-square tests of independence. Where expected cell counts were small, Fisher’s exact test was used for 2 × 2 contingency tables, and the Fisher–Freeman–Halton exact test with Monte Carlo significance was used for larger contingency tables. Statistical significance was set at *p* < .05.

Intervention effects were examined using linear mixed-effects models including fixed effects for group, time, and their interaction, with a random intercept for participant, allowing inclusion of all available data under an intention-to-treat framework. Group and time were specified as categorical fixed factors in all models; no covariates were included in the primary analysis to preserve the randomised comparison. Linear mixed-effects models were fitted with a random intercept for participant and an autoregressive [AR(1)] covariance structure for repeated measures over time. This structure was selected to appropriately model within-participant correlations and to ensure model convergence in the presence of incomplete follow-up data.

Exploratory multiple linear regression was conducted to examine whether selected baseline demographic and clinical characteristics were associated with treatment response. Treatment response was operationalised as change in TFI score from baseline to 12-week follow-up, calculated as baseline minus follow-up score so that positive values indicated improvement. The model included group allocation, baseline TFI score, age, tinnitus duration, concurrent hyperacusis, and baseline DASS-21 anxiety as predictors. Predictors were selected a priori based on clinical relevance and observed differences in attrition analyses. Due to the sample size, variables with sparse response categories were not included in the model.

Exploratory program-usage analyses were conducted within the intervention group to examine whether engagement with Tune Out was associated with differential change in outcomes over time. Program usage was operationalised as total duration of program use and dichotomised as less than 2 hours versus more than 2 hours. Separate linear mixed-effects models were fitted for tinnitus severity, tinnitus handicap, and psychological distress. These models included fixed effects for time, program use group, and their interaction, with a random intercept for participant and an AR(1) covariance structure for repeated measures. The Time × Program Use interaction was used to assess whether outcome trajectories differed between participants with lower and higher program use. These analyses were considered exploratory.

Exploratory analyses examined whether baseline self-efficacy was associated with program engagement and treatment response within the intervention group. Baseline GSE scores were correlated with total program use duration using Spearman’s rho due to the skewed distribution of usage data. Additional correlations examined associations between baseline GSE scores and changes in tinnitus severity, tinnitus handicap, and psychological symptoms from baseline to follow-up, with positive change scores indicating improvement.

## RESULTS

### Participant flow and baseline characteristics

Of the 115 individuals assessed for eligibility, 88 were randomised: 43 to the intervention group and 45 to the waitlist control group (Figure 1). Twenty-seven individuals were excluded prior to randomisation, most commonly because they declined to participate (n = 25). All randomised participants received their allocated condition. By the final follow-up, 17 participants in the intervention group and eight participants in the waitlist control group had withdrawn or were lost to follow-up. The primary outcome analysis therefore included 63 participants: 26 in the intervention group and 37 in the waitlist control group.

Baseline demographic and clinical characteristics by randomised group are presented in Table 1. Participants had a mean age of 62.53 years (SD = 12.60), and most reported constant tinnitus (95.45%). Tinnitus was most commonly perceived bilaterally (69.32%), and the mean duration of tinnitus was 14.32 years (SD = 15.06). Almost half of participants reported concurrent hyperacusis (47.73%). At baseline, the mean TFI score was 57.54 (SD = 25.18) and the mean THI score was 43.86 (SD = 26.75). No statistically significant differences were identified between the intervention and waitlist control groups across baseline demographic, tinnitus-related, psychological, self-efficacy, or technology-use variables.

**Table 1.** Baseline Demographic and Clinical Characteristics by Randomised Group.

|  | Total<br>N=88<br>n(%) or M(SD) | Intervention<br>n=43<br>n(%) or M(SD) | Control<br>n=45<br>n(%) or M(SD) | Group<br>comparison<br>p |
| --- | --- | --- | --- | --- |
| Age M(SD) | 62.53 (12.60) | 61.74 (12.56) | 63.29 (12.74) | .568<sup>a</sup> |
| Gender Female | 46 (52.27) | 25 (58.14) | 24 (53.33) |  |
| Male | 42 (47.72) | 18 (41.86) | 21 (46.67) | .281<sup>b</sup> |
| Highest level of education |  |  |  |  |
| Post-graduate | 19 (21.84) | 9 (20.93) | 10 (22.73) |  |
| Undergraduate | 41 (47.13) | 24 (55.81) | 17 (38.64) |  |
| Secondary school | 24 (27.59) | 8 (18.60) | 16 (36.36) |  |
| Other | 1 (1.15) | 0 | 1 (2.27) |  |
| Prefer not to say | 2 (2.30) | 2 (4.65) | 0 | .115<sup>c</sup> |
| Duration of tinnitus (years) M(SD) | 14.32 (15.06) | 15.89 (17.47) | 12.83 (12.34) | .976<sup>d</sup> |
| Frequency of tinnitus Constant | 84 (95.45) | 41 (95.35) | 43 (95.56) |  |
| Intermittent | 4 (4.54) | 2 (2.27) | 2 (4.44) | .963<sup>b</sup> |
| Location of tinnitus Right ear | 12 (13.64) | 6 (13.95) | 6 (13.33) |  |
| Left ear | 14 (15.91) | 6 (13.95) | 8 (17.78) |  |
| Central / both ears | 61 (69.32) | 31 (72.09) | 30 (66.67) | .865<sup>b</sup> |
| Concurrent hyperacusis | 42 (47.73) | 22 (51.16) | 20 (44.44) | .528<sup>b</sup> |
| History of sudden hearing loss | 14 (15.91) | 8 (18.60) | 6 (13.33) | .499<sup>b</sup> |
| History of noise exposure | 61 (69.32) | 31 (72.09) | 30 (68.18) | .690<sup>b</sup> |
| Providers seen for hearing or tinnitus |  |  |  |  |
| Ear nose and throat specialist (ENT) | 54 (61.36) | 23 (53.49) | 31 (68.89) | .138<sup>b</sup> |
| Audiologist | 70 (79.55) | 35 (81.40) | 35 (77.78) | .674<sup>b</sup> |
| General practitioner (GP) | 64 (72.73) | 31 (72.09) | 30 (68.18) | .896<sup>b</sup> |
| Other | 7 (7.95) | 6 (13.95) | 1 (2.27) | .055<sup>e</sup> |
| Management received for hearing or tinnitus |  |  |  |  |
| Hearing aid | 33 (37.50) | 15 (34.88) | 18 (40.0) | .620<sup>b</sup> |
| Cochlear implant | 2 (2.30) | 2 (2.27) | 0 | .236<sup>e</sup> |
| Assistive listening device | 5 (5.68) | 3 (6.98) | 2 (4.44) | .673<sup>e</sup> |
| Communication training / education | 4 (4.54) | 1 (2.33) | 3 (6.67) | .617<sup>e</sup> |
| Tinnitus management | 19 (21.59) | 8 (18.60) | 11 (24.44) | .506<sup>b</sup> |
| None | 37 (42.05) | 19 (44.19) | 18 (40.0) | .691<sup>b</sup> |
| Previous diagnosis of mental disorder |  |  |  |  |
| Depression | 16 (18.18) | 7 (16.28) | 9 (20.0) | .651<sup>b</sup> |
| Anxiety | 23 (26.14) | 11 (25.58) | 12 (26.67) | .908<sup>b</sup> |
| Post traumatic stress disorder (PTSD) | 7 (7.95) | 4 (9.30) | 3 (6.67) | .710<sup>e</sup> |
| Self-rated ability to use technology |  |  |  |  |
| Computers Excellent | 43 (48.86) | 24 (55.81) | 19 (42.22) |  |
| Good | 41 (47.13) | 16 (37.21) | 25 (55.56) |  |
| Poor | 4 (4.54) | 3 (6.98) | 1 (2.27) | .185<sup>c</sup> |
| Smart devices Excellent | 43 (48.86) | 25 (58.14) | 18 (40.0) |  |
| Good | 41 (47.13) | 15 (34.88) | 26 (60.47) |  |
| Poor | 4 (4.54) | 3 (6.98) | 1 (2.27) | .057<sup>c</sup> |
| Internet Excellent | 48 (54.55) | 27 (62.79) | 21 (46.67) |  |
| Good | 39 (44.32) | 15 (34.88) | 24 (53.33) |  |
| Poor | 1 (1.15) | 1 (2.33) | 0 | .109<sup>c</sup> |
| Tinnitus Functional Index (TFI) | 57.54 (25.18) | 61.27 (26.39) | 53.98 (23.73) | .171<sup>d</sup> |
| Intrusiveness sub-scale | 70.15 (24.57) | 71.71 (25.68) | 68.67 (23.65) | .454<sup>d</sup> |
| Sense of control sub-scale | 69.85 (25.11) | 74.65 (24.67) | 65.26 (24.93) | .051<sup>d</sup> |
| Cognitive sub-scale | 50.80 (29.21) | 54.57 (30.44) | 47.19 (27.84) | .219<sup>d</sup> |
| Sleep sub-scale | 59.02 (33.16) | 62.87 (34.20) | 55.33 (32.08) | .300<sup>d</sup> |
| Auditory sub-scale | 55.64 (32.83) | 55.04 (31.69) | 56.22 (34.23) | .957<sup>d</sup> |
| Relaxation sub-scale | 67.39 (32.13) | 73.10 (33.20) | 61.93 (30.43) | .072<sup>d</sup> |
| Quality of life sub-scale | 45.99 (30.11) | 50.87 (31.90) | 41.33 (27.86) | .235<sup>d</sup> |
| Emotional sub-scale | 45.34 (31.50) | 50.78 (34.94) | 40.15 (27.20) | .267<sup>d</sup> |
| Tinnitus Handicap Inventory (THI) | 43.86 (26.75) | 44.88 (27.65) | 42.89 (26.13) | .767<sup>d</sup> |
| Functional sub-scale | 20.25 (12.18) | 20.84 (12.35) | 19.69 (12.13) | .601<sup>d</sup> |
| Emotional sub-scale | 13.46 (10.86) | 13.63 (11.24) | 13.29 (10.62) | .970<sup>d</sup> |
| Catastrophic sub-scale | 10.16 (4.97) | 10.42 (5.23) | 9.91 (4.75) | .730<sup>d</sup> |
| Depression Anxiety and Stress Scale 21 item | 23.09 (19.73) | 21.86 (19.90) | 24.27 (19.72) | .272<sup>d</sup> |
| Depression subscale | 7.02 (8.25) | 6.74 (7.60) | 7.29 (8.92) | .657<sup>d</sup> |
| Anxiety subscale | 5.61 (5.61) | 5.02 (5.75) | 6.18 (5.49) | .147<sup>d</sup> |
| Stress subscale | 10.46 (7.68) | 10.09 (7.92) | 10.80 (7.52) | .557<sup>d</sup> |
| General self-efficacy scale (GSE-10) | 31.51 (4.45) | 31.37 (4.43) | 31.64 (4.51) | .471<sup>d</sup> |

<sup>a</sup>Independent samples t-test
<sup>b</sup>Pearson's chi-square test of independence
<sup>c</sup>Fisher-Freeman-Halton Exact Test (Monte Carlo Significance)
<sup>d</sup>Mann-Whitney U
<sup>e</sup>Fisher's Exact Test

Characteristics of participants who completed at least one follow-up assessment and those who dropped out are presented in Table 2. Completers and dropouts were similar across most baseline characteristics, including baseline tinnitus severity and tinnitus handicap. However, participants who dropped out reported a significantly shorter duration of tinnitus and higher baseline anxiety symptoms than those who completed at least one follow-up assessment.

**Table 2.** Baseline Demographic and Clinical Characteristics by Completion Status.

|  | Total<br>N=88 | Completed at<br>least one<br>follow-up n=74 | Dropped out<br>n=14 | Group<br>comparison<br>*p* |
| --- | --- | --- | --- | --- |
|  | n(%) or M(SD) | n(%) or M(SD) | n(%) or M(SD) |  |
| Age M(SD) | 62.53 (12.60) | 63.47 (12.91) | 57.57 (9.74) | .108<sup>a</sup> |
| Gender |  |  |  |  |
| Female | 46 (52.3) | 37 (50.0) | 9 (64.3) |  |
| Male | 42 (47.7) | 37 (50.0) | 5 (35.7) | .326<sup>b</sup> |
| Highest level of education |  |  |  |  |
| Post-graduate | 19 (21.8) | 16 (21.6) | 3 (23.1) |  |
| Undergraduate | 41 (47.1) | 36 (48.6) | 5 (38.5) |  |
| Secondary school | 24 (27.6) | 20 (27.0) | 4 (30.8) |  |
| Other | 1 (1.2) | 1 (1.4) | 0 |  |
| Prefer not to say | 2 (2.3) | 1 (1.4) | 1 (7.7) | .506<sup>c</sup> |
| Duration of tinnitus (years) M(SD) | 14.32 (15.06) | 15.92 (15.30) | 6.13 (10.86) | .008<sup>d</sup> |
| Frequency of tinnitus |  |  |  |  |
| Constant | 84 (95.5) | 72 (97.3) | 12 (85.7) |  |
| Intermittent | 4 (4.5) | 2 (2.7) | 2 (14.3) | .117<sup>b</sup> |
| Location of tinnitus |  |  |  |  |
| Right ear | 12 (13.6) | 10 (13.7) | 2 (14.3) |  |
| Left ear | 14 (15.9) | 9 (12.3) | 5 (35.7) |  |
| Central / both ears | 61 (69.3) | 54 (74.0) | 7 (50.0) | .072<sup>d</sup> |
| Concurrent hyperacusis | 42 (47.7) | 37 (50.0) | 5 (35.7) | .326<sup>b</sup> |
| History of sudden hearing loss | 14 (15.9) | 12 (16.2) | 2 (14.3) | .856<sup>b</sup> |
| History of noise exposure | 61 (69.3) | 54 (74.0) | 7 (50.0) | .073<sup>b</sup> |
| Providers seen for hearing or tinnitus |  |  |  |  |
| Ear nose and throat specialist (ENT) | 54 (61.4) | 47 (63.5) | 7 (50.0) | .341<sup>b</sup> |
| Audiologist | 70 (79.6) | 59 (79.7) | 11 (78.6) | .922<sup>b</sup> |
| General practitioner (GP) | 64 (72.7) | 53 (71.6) | 11 (78.6) | .592<sup>b</sup> |
| Other | 7 (8.0) | 6 (8.1) | 1 (7.1) | .903<sup>b</sup> |
| Management received for hearing or tinnitus |  |  |  |  |
| Hearing aid | 33 (37.5) | 30 (40.5) | 3 (21.4) | .176<sup>b</sup> |
| Cochlear implant | 2 (2.3) | 2 (2.7) | 0 | .534<sup>e</sup> |
| Assistive listening device | 5 (5.7) | 5 (6.8) | 0 | .317<sup>e</sup> |
| Communication training / education | 4 (4.54) | 3 (4.1) | 1 (7.1) | .611<sup>e</sup> |
| Tinnitus management | 19 (21.6) | 18 (24.3) | 1 (7.1) | .152<sup>b</sup> |
| None | 37 (42.1) | 29 (39.2) | 8 (57.1) | .212<sup>b</sup> |
| Previous diagnosis of mental disorder |  |  |  |  |
| Depression | 16 (18.2) | 14 (18.9) | 2 (14.3) | .680<sup>b</sup> |
| Anxiety | 23 (26.1) | 20 (27.0) | 3 (21.4) | .662<sup>b</sup> |
| Post traumatic stress disorder (PTSD) | 7 (8.0) | 7 (9.5) | 0 | .591<sup>e</sup> |
| Self-rated ability to use technology |  |  |  |  |
| Computers |  |  |  |  |
| Excellent | 43 (48.9) | 35 (47.3) | 8 (57.1) |  |
| Good | 41 (47.1) | 35 (47.3) | 6 (42.9) |  |
| Poor | 4 (4.5) | 4 (5.4) | 0 | .594<sup>c</sup> |
| Smart devices |  |  |  |  |
| Excellent | 43 (48.9) | 35 (47.3) | 8 (57.1) |  |
| Good | 41 (47.1) | 36 (48.6) | 5 (35.7) |  |
| Poor | 4 (4.5) | 3 (4.1) | 1 (7.1) | .637<sup>c</sup> |
| Internet |  |  |  |  |
| Excellent | 48 (54.6) | 48 (54.5) | 11 (78.6) |  |
| Good | 39 (44.3) | 39 (44.3) | 3 (21.4) |  |
| Poor | 1 (1.2) | 1 (1.1) | 0 | .141<sup>c</sup> |
| Tinnitus Functional Index (TFI) | 57.54 (25.18) | 57.63 (24.29) | 57.06 (30.51) | .927<sup>d</sup> |
| Intrusiveness sub-scale | 70.15 (24.57) | 70.95 (24.06) | 65.95 (27.68) | .468<sup>d</sup> |
| Sense of control sub-scale | 69.85 (25.11) | 69.41 (24.66) | 72.14 (28.24) | .775<sup>d</sup> |
| Cognitive sub-scale | 50.80 (29.21) | 51.13 (29.08) | 49.05 (30.95) | .766<sup>d</sup> |
| Sleep sub-scale | 59.02 (33.16) | 58.87 (33.38) | 59.76 (33.14) | .950<sup>d</sup> |
| Auditory sub-scale | 55.64 (32.83) | 56.76 (32.48) | 49.76 (35.29) | .541<sup>d</sup> |
| Relaxation sub-scale | 67.39 (32.13) | 68.24 (30.69) | 62.86 (39.91) | .891<sup>d</sup> |
| Quality of life sub-scale | 45.99 (30.11) | 45.91 (30.02) | 46.43 (31.72) | .923<sup>d</sup> |
| Emotional sub-scale | 45.34 (31.50) | 43.69 (30.63) | 54.05 (35.71) | .404<sup>d</sup> |
| Tinnitus Handicap Inventory (THI) | 43.86 (26.75) | 43.30 (25.62) | 46.86 (33.04) | .797<sup>d</sup> |
| Functional sub-scale | 20.25 (12.18) | 20.14 (11.77) | 20.86 (14.67) | .762<sup>d</sup> |
| Emotional sub-scale | 13.46 (10.86) | 13.14 (10.61) | 15.14 (12.42) | .697<sup>d</sup> |
| Catastrophic sub-scale | 10.16 (4.97) | 10.03 (4.68) | 10.86 (6.46) | .589<sup>d</sup> |
| Depression Anxiety and Stress Scale 21 item | 23.09 (19.73) | 22.05 (19.66) | 28.57 (19.91) | .169<sup>d</sup> |
| Depression subscale | 7.02 (8.25) | 6.68 (8.30) | 8.86 (8.07) | .278<sup>d</sup> |
| Anxiety subscale | 5.61 (5.61) | 5.14 (5.58) | 8.14 (5.29) | .032<sup>d</sup> |
| Stress subscale | 10.46 (7.68) | 10.24 (7.68) | 11.57 (7.89) | .362<sup>d</sup> |
| General self-efficacy scale (GSE-10) | 31.51 (4.45) | 31.31 (4.55) | 32.57 (3.84) | .319<sup>d</sup> |

<sup>a</sup>Independent samples t-test
<sup>b</sup>Pearson's chi-square test of independence
<sup>c</sup>Fisher-Freeman-Halton Exact Test (Monte Carlo Significance)
<sup>d</sup>Mann-Whitney U
<sup>e</sup>Fisher's Exact Test

### Primary and secondary efficacy outcomes

Descriptive outcome data across baseline, 6-week follow-up, and 12-week follow-up are presented in Table 3, and linear mixed-effects model results are presented in Table 4. For the primary outcome, there was a significant Group × Time interaction for TFI total score, *F*(2, 102.57) = 5.95, *p* = .004, partial η² = .104, indicating that change in tinnitus severity over time differed between groups. The intervention group showed a reduction in mean TFI scores from baseline to 6 weeks, which was maintained at 12 weeks. In contrast, the control group showed an increase in TFI scores at 6 weeks and remained above baseline at 12 weeks (Figure 2). Significant Group × Time interactions were also observed across several TFI subscales, including intrusiveness, sense of control, auditory difficulties, relaxation, quality of life, and emotional distress, while the cognitive and sleep subscales were not significant (See Supplementary Material 1).

**Figure 2.**
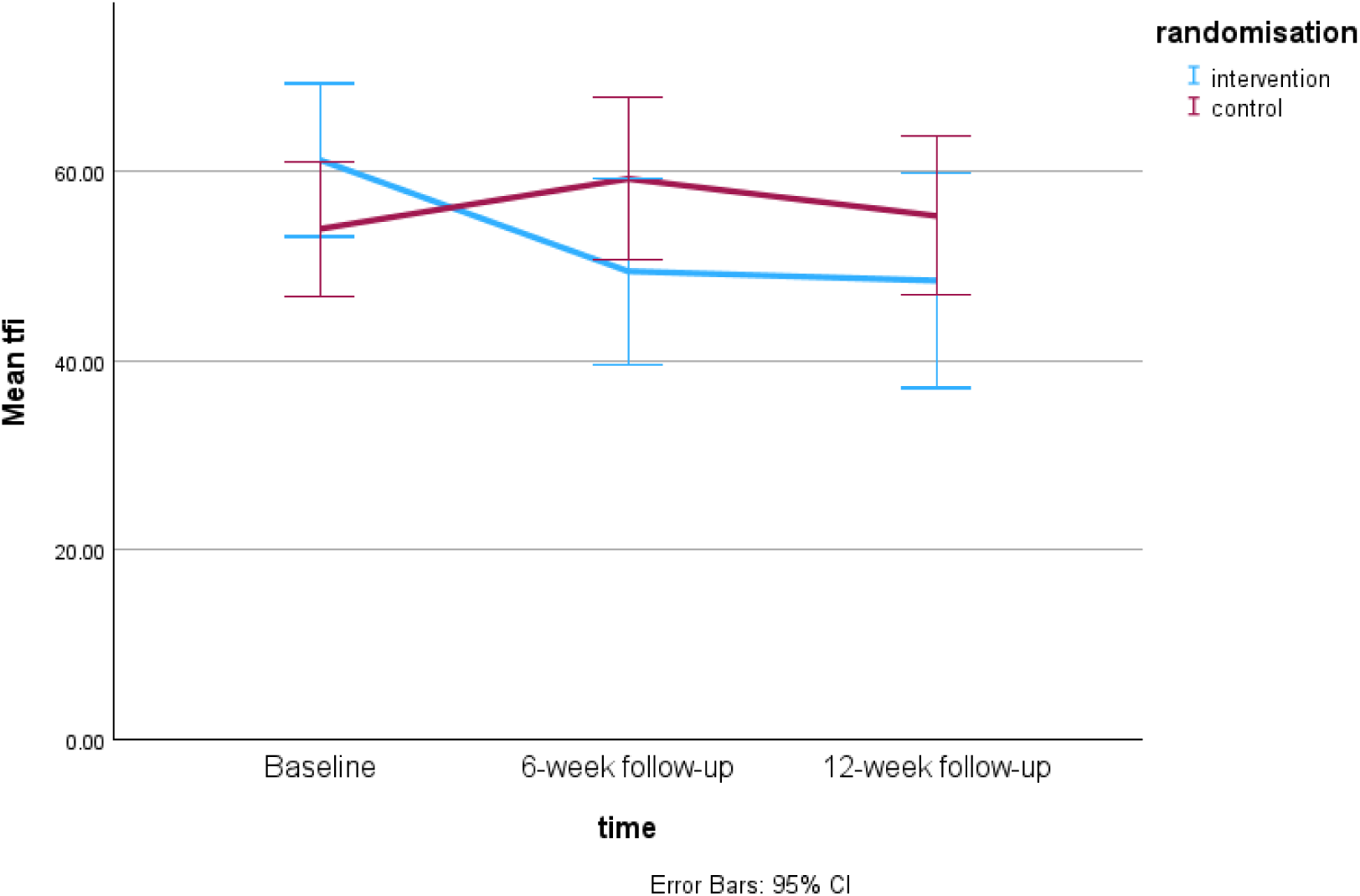
Mean TFI scores across time by randomised group.

**Table 3.** Descriptive Outcome Scores by Group and Time Point.

|  | Baseline |  | 6-week follow up |  | 12-week follow up |  |
| --- | --- | --- | --- | --- | --- | --- |
|  | Intervention<br>M(SD) | Control<br>M(SD) | Intervention<br>M(SD) | Control<br>M(SD) | Intervention<br>M(SD) | Control<br>M(SD) |
| TFI | 61.27 (26.39) | 53.98 (23.73) | 49.49 (26.90) | 59.28 (26.47) | 48.51 (28.16) | 55.35 (25.17) |
| THI | 44.88 (27.65) | 42.89 (26.13) | 35.16 (25.63) | 49.23 (27.85) | 40.06 (27.45) | 45.05 (26.40) |
| DASS-21 Total | 21.86 (19.90) | 24.27 (19.72) | 18.90 (16.35) | 28.31 (23.71) | 20.95 (18.53) | 25.53 (20.74) |
| DASS-21 Depression | 6.74 (7.60) | 7.29 (8.92) | 6.90 (7.64) | 7.80 (9.50) | 7.13 (7.89) | 7.46 (9.12) |
| DASS-21 Anxiety | 5.02 (5.75) | 6.18 (5.49) | 3.55 (4.58) | 7.28 (6.70) | 4.50 (5.31) | 6.30 (5.90) |
| DASS-21 Stress | 10.09 (7.92) | 10.80 (7.52) | 8.45 (6.83) | 13.23 (9.33) | 9.33 (7.25) | 11.77 (7.92) |

**Note.** Values are presented as *M* (SD). TFI = Tinnitus Functional Index; THI = Tinnitus Handicap Inventory; DASS-21 = Depression Anxiety Stress Scale–21 items

**Table 4.** Fixed Effects from Linear Mixed-Effects Models for Primary and Secondary Outcomes.

| | Effect | F (df<sub>1</sub>,df<sub>2</sub>) | p | Partial $\eta^2$ |
| --- | --- | --- | --- | --- |
| TFI | Group | F(1,86.32) = 0.003 | .958 | <.001 |
|  | Time | F(2,102.57) = 1.75 | .179 | .033 |
|  | Group × Time | F(2,102.57) = 5.95 | **.004** | .104 |
| THI | Group | F(1,85.56) = 0.18 | .670 | .002 |
|  | Time | F(2,106.76) = 1.07 | .349 | .020 |
|  | Group × Time | F(2,106.76) = 4.12 | **.019** | .072 |
| DASS-21 depression subscale | Group | F(1,87.68) = 0.06 | .808 | .001 |
|  | Time | F(2,99.66) = 2.08 | .130 | .040 |
|  | Group × Time | F(2,99.66) = 1.72 | .184 | .033 |
| DASS-21 anxiety subscale | Group | F(1,86.24) = 1.16 | .285 | .013 |
|  | Time | F(2,116.85) = 0.07 | .930 | .001 |
|  | Group × Time | F(2,116.85) = 3.63 | **.030** | .059 |
| DASS-21 stress subscale | Group | F(1,85.85) = 1.72 | .194 | .020 |
|  | Time | F(2,108.79) = 0.70 | .501 | .013 |
|  | Group × Time | F(2,108.79) = 2.84 | .063 | .050 |

**Note.** TFI = Tinnitus Functional Index; THI = Tinnitus Handicap Inventory; DASS-21 = Depression Anxiety Stress Scale–21 items. Group refers to randomised allocation; Time refers to baseline, 6-week follow-up, and 12-week follow-up. Partial $\eta^2$ values were calculated from the F statistic and associated degrees of freedom.

For tinnitus handicap, there was also a significant Group × Time interaction for THI total score, *F*(2, 106.76) = 4.12, *p* = .019, partial η² = .072. Mean THI scores decreased in the intervention group at 6 weeks, with partial maintenance at 12 weeks, whereas scores in the control group increased at 6 weeks and remained above baseline at 12 weeks. At the subscale level, significant interactions were observed for the emotional and catastrophic subscales, but not the functional subscale.

For psychological outcomes, the Group × Time interaction for DASS-21 total score did not reach statistical significance, *F*(2, 113.50) = 2.74, *p* = .069, partial η² = .046. However, a significant interaction was observed for the DASS-21 anxiety subscale, *F*(2, 116.85) = 3.63, *p* = .030, partial η² = .059. Interactions for the depression and stress subscales were not statistically significant. This pattern suggests that the intervention effect was more evident for tinnitus-related outcomes than for broader psychological distress.

Overall, the pattern of results suggested greater reductions in tinnitus severity and tinnitus handicap over time in the intervention group compared with waitlist control, with the clearest effects observed for TFI total score and several tinnitus-related subdomains. Effects on broader psychological symptoms were less consistent, although anxiety symptoms showed a significant differential change over time.

### Responder analyses

Responder analyses provided additional evidence of improvement in tinnitus severity. At 12 weeks, 69.2% of participants in the intervention group showed improvement in TFI scores compared with 40.5% in the control group, χ²(1, *N* = 63) = 5.04, *p* = .025, OR = 3.30, 95% CI [1.14, 9.52]. Clinically meaningful improvement in tinnitus severity, defined as a ≥13-point reduction in TFI, was also more common in the intervention group than the control group, 23.1% versus 5.4%, χ²(1, *N* = 63) = 4.30, *p* = .038; however, the confidence interval around the odds ratio was wide and included 1, OR = 5.25, 95% CI [0.97, 28.57]. For THI and DASS-21, the proportion of participants showing improvement did not differ significantly between groups.

### Exploratory and sensitivity analyses

An exploratory multiple linear regression examined whether group allocation predicted TFI improvement from baseline to 12 weeks after adjustment for baseline TFI, age, tinnitus duration, concurrent hyperacusis, and baseline DASS-21 anxiety. The overall model was not statistically significant, *F*(6, 54) = 1.76, *p* = .126, *R*² = .163, adjusted *R*² = .070. However, group allocation was a significant independent predictor of TFI improvement, *B* = 9.48, 95% CI [0.52, 18.44], *p* = .039, indicating that participants in the intervention group showed approximately 9.5 points greater improvement than those in the control group after adjustment. No other predictors were statistically significant, and collinearity statistics were acceptable.

Sensitivity analyses excluding intervention participants with no recorded program use supported the primary findings. Group × Time interactions remained significant for TFI, *F*(2, 95.55) = 7.22, *p* = .001, partial η² = .131, and THI, *F*(2, 100.83) = 4.72, *p* = .011, partial η² = .086. Significant interactions were also observed for DASS-21 total score, *F*(2, 105.48) = 4.31, *p* = .016, partial η² = .076, anxiety, *F*(2, 108.97) = 4.81, *p* = .010, partial η² = .081, and stress, *F*(2, 101.99) = 3.53, *p* = .033, partial η² = .065, but not depression, *F*(2, 91.77) = 2.26, *p* = .110, partial η² = .047. As these analyses were based on treatment received, they should be interpreted as supportive and exploratory rather than confirmatory.

### Program engagement and self-efficacy

Program usage data indicated substantial variability in engagement among participants allocated to the intervention group. Mean program use was 8.45 hours (SD = 19.11), although the distribution was highly skewed, with a median of 1.10 hours and an interquartile range of 7.50 hours. Recorded usage ranged from 0 to 89 hours, with 25.6% of participants randomised to the intervention not using the program at all. This pattern suggests that while some participants engaged with the program extensively, typical use was relatively low.

Exploratory analyses within the intervention group examined whether program usage was associated with differential change in outcomes over time. Program usage was dichotomised as less than 2 hours versus 2 hours or more of use. Program usage was associated with change in tinnitus severity, with a significant Time × Program Use interaction for TFI scores, *F*(2, 40.58) = 3.35, *p* = .045, partial η² = .14. This indicated that TFI trajectories differed between participants with lower and higher program use. There was also a significant main effect of time for TFI, *F*(2, 40.58) = 5.64, *p* = .007, but no main effect of program use duration, *F*(1, 41.42) = 0.21, *p* = .648.

In contrast, program usage was not associated with differential change in tinnitus handicap or psychological distress. For THI, there was a significant main effect of time, *F*(2, 47.80) = 3.24, *p* = .048, but no main effect of program use duration, *F*(1, 40.77) = 0.06, *p* = .806, and no Time × Program Use interaction, *F*(2, 47.80) = 0.61, *p* = .548. For DASS-21, there were no significant effects of time, program use duration, or their interaction. Overall, these exploratory findings suggest that greater program use was associated with a different pattern of change in tinnitus severity, but not tinnitus handicap or psychological distress.

Baseline self-efficacy was not significantly associated with program engagement or treatment response within the intervention group. Specifically, baseline GSE scores were not associated with total program use duration, *r*s = −.032, *p* = .840, or change in TFI, *r*s = .200, *p* = .328, THI, *r*s = .169, *p* = .408, or DASS-21 scores, *r*s = −.057, *p* = .780. These findings suggest that general self-efficacy did not predict engagement with Tune Out or improvement in tinnitus-related or psychological outcomes in this sample.

### Program usability

Usability of Tune Out was assessed using the System Usability Scale at follow-up. Among participants who completed the usability measure, the mean SUS score was 73.13 (SD = 18.29; range = 40.00–97.50; *N* = 24), indicating generally good perceived usability. This exceeds the commonly cited SUS benchmark of 68, suggesting that Tune Out was perceived as having above-average usability among respondents.

## DISCUSSION

This study evaluated the efficacy, engagement, and usability of Tune Out, an unguided, self-paced online tinnitus management program. Overall, the findings provide preliminary support for Tune Out as an accessible iCBT-based intervention for tinnitus. Participants allocated to the intervention demonstrated greater improvements in tinnitus severity compared with those allocated to the waitlist control group, with improvements maintained at 12-week follow-up. Improvements were also observed for tinnitus handicap, particularly in emotional and catastrophic responses to tinnitus, suggesting that the program may help reduce the perceived burden and distress associated with tinnitus. Effects on broader psychological symptoms were less consistent, although anxiety showed evidence of differential improvement over time. Program engagement was highly variable and generally low, but exploratory analyses suggested that participants who used the program for longer showed more favourable changes in tinnitus severity. Usability ratings were above commonly accepted benchmarks, indicating that participants who completed the usability measure generally perceived Tune Out as usable and acceptable.

The strongest evidence of benefit was observed for tinnitus severity, as measured by the TFI. The intervention group demonstrated reductions in TFI scores at 6 weeks that were maintained at 12 weeks, while the waitlist control group showed less favourable change over the same period. Significant intervention effects were also observed across several TFI subdomains, including intrusiveness, sense of control, auditory difficulties, relaxation, quality of life, and emotional distress. This pattern is consistent with the intended mechanism of CBT-based tinnitus interventions, which do not aim to eliminate the tinnitus percept itself, but instead seek to reduce its intrusiveness, emotional salience, and functional impact (Jun & Park, 2013). The significant effects for sense of control, relaxation, quality of life, and emotional distress are particularly relevant, as these domains align closely with the program’s focus on cognitive reframing, behavioural strategies, arousal reduction, and acceptance-based coping.

The observed improvement in tinnitus severity is consistent with previous evidence supporting internet-delivered CBT for tinnitus. Prior reviews and trials have suggested that iCBT can reduce tinnitus distress and related functional impact, and the current findings extend this literature by evaluating a fully unguided and self-paced program (Beukes et al., 2019; Demoen et al., 2023; Fuller et al., 2020; Sia et al., 2024). This is important because unguided digital interventions may offer a scalable option for people who cannot access tinnitus-specific psychological care due to geographic, financial, workforce, or service availability barriers (Maes et al., 2014; Patel et al., 2022). However, unguided interventions also place greater responsibility on participants to initiate, continue, and apply program content independently.

Responder analyses provided further support for the impact of Tune Out on tinnitus severity, although the proportion achieving clinically meaningful change was lower than reported in some previous iCBT studies. In the present study, 23.1% of participants in the intervention group achieved a clinically meaningful reduction in tinnitus severity, defined as a ≥13-point reduction in TFI (Henry et al., 2024). This is lower than the proportions reported in other 6–8-week tinnitus iCBT studies, including Andersson et al. (2002), who reported clinically significant improvement in 31% of participants using unguided iCBT, and Kaldo et al. (2008) and Beukes et al. (2018), who reported improvement rates of 38% and 51%, respectively, in guided iCBT interventions. One possible explanation for the lower responder rate in the present study is the variability in program engagement, as one-quarter of participants allocated to the intervention recorded no program use. These findings suggest that Tune Out may be beneficial for some users, but that unguided access alone may not be sufficient to ensure clinically meaningful improvement for a substantial proportion of participants.

Improvements in tinnitus handicap were also observed, although the pattern was less consistent than for tinnitus severity. The Group × Time interaction was significant for THI total score, and subscale analyses indicated effects for the emotional and catastrophic subscales, but not the functional subscale. This may suggest that Tune Out primarily influenced participants’ emotional and cognitive responses to tinnitus rather than broader functional limitations. This interpretation is consistent with the CBT and ACT components of the program, which focus on changing the meaning, threat value, catastrophising, and behavioural response to tinnitus (Gellatly & Beck, 2016). However, responder analyses for clinically meaningful change in THI did not clearly favour the intervention group, suggesting that while group-level improvements were observed, these did not consistently translate into a higher proportion of participants achieving clinically meaningful improvement in tinnitus handicap.

Effects on broader psychological symptoms were more modest. The Group × Time interaction for DASS-21 total score did not reach statistical significance in the primary analysis, although anxiety symptoms showed significant differential change over time. This may indicate that Tune Out had some impact on tinnitus-related worry, arousal, or threat monitoring, but was not sufficient to produce broader improvements in depression or stress symptoms. This finding is not unexpected, as Tune Out is a tinnitus management program rather than a mental health intervention, and several other studies showed similar results (Aazh et al., 2024; Fuller et al., 2020). Participants with more complex or persistent psychological distress may require additional guided psychological support, stepped-care pathways, or referral to appropriately trained mental health professionals alongside digital tinnitus management.

Program engagement was highly variable and typical use was relatively low. Although mean program use was influenced by a small number of high-use participants, the median use was low and approximately one-quarter of participants allocated to the intervention recorded no program use. This highlights a central challenge for unguided digital interventions, that accessibility alone does not guarantee uptake or sustained engagement. This pattern is consistent with broader evidence on online psychological interventions, where adherence is recognised as a key challenge. In a systematic review of predictors of adherence to online psychological interventions, Beatty and Binnion (2016) found that guidance or support, sufficient time, higher treatment expectancy, and higher satisfaction with intervention content were associated with greater adherence, while baseline symptom severity and education were not consistently related to adherence. These findings provide a useful context for the present study, where low and variable engagement may have been influenced by factors not captured in the current dataset, such as time availability, treatment expectations, or perceived relevance of program content.

The engagement findings also raise the question of whether some level of guidance may improve outcomes. Guided iCBT, which includes brief support from a psychologist, audiologist, or trained clinician, appears particularly promising. Several studies suggest that internet-based therapies may be more effective when some clinician guidance is provided, although this support does not necessarily require full-length consultations (Andersson, 2015; Demoen et al., 2023; Sattel et al., 2025). Guided iCBT has generally been found to reduce tinnitus impact and distress (Andersson, 2016; Demoen et al., 2023; Karyotaki et al., 2021), and may offer advantages for supporting adherence, troubleshooting barriers, and encouraging continued use. In this context, Tune Out may be best understood as a scalable low-intensity intervention, but future research should examine whether optional brief guidance, automated prompts, or stepped-care support could improve engagement and increase the proportion of participants achieving clinically meaningful benefit.

Sensitivity analyses excluding intervention participants with no recorded program use supported the primary findings. The intervention effects on tinnitus severity and tinnitus handicap remained significant, and additional effects were observed for DASS-21 total score, anxiety, and stress. These findings suggest that Tune Out may have greater effects among participants who actually access the program. However, these analyses were based on treatment received rather than randomised allocation alone and therefore do not preserve the full benefits of randomisation. As such, they should be interpreted as supportive and exploratory rather than confirmatory.

Although self-efficacy was conceptually relevant to unguided intervention engagement, baseline general self-efficacy was not associated with program use or treatment response. This may indicate that general self-efficacy was not a key determinant of engagement or outcomes in this sample. However, it is also possible that the General Self-Efficacy Scale was not sufficiently specific to capture confidence in managing tinnitus, using digital health tools, or persisting with self-guided CBT-based activities. Specific scales such as the Internet Intervention Adherence Self-Efficacy Scale (IIASES) (Smoktunowicz et al., 2024) may be a more sensitive predictor of engagement with unguided tinnitus interventions and should be considered in future studies.

Participants who completed the usability measure rated Tune Out favourably, with mean SUS scores above the commonly cited benchmark for acceptable usability (Sauro & Lewis, 2016). This suggests that, among respondents, the program was generally perceived as usable. However, favourable usability ratings should be interpreted alongside the low and variable engagement data. A program may be easy to use for those who access it, while still requiring additional strategies to support uptake, adherence, and completion. Future iterations of Tune Out could consider incorporating methods of promoting engagement (Saleem et al., 2021), such as automated reminders, onboarding support, progress feedback or optional brief clinician contact.

### Strengths, limitations and future research

This study has several strengths. It used a randomised controlled design, included a waitlist control group, and assessed outcomes across multiple time points. The study also used validated measures of tinnitus severity, tinnitus handicap, psychological symptoms, self-efficacy, and usability. In addition, the inclusion of program usage data allowed exploration of engagement patterns, which is particularly important for unguided digital interventions. The study therefore provides useful evidence not only about whether Tune Out may be effective, but also about how participants engage with this type of self-guided intervention in practice.

Several limitations should also be acknowledged. First, attrition was higher in the intervention group than the waitlist control group, which may have introduced bias despite the use of mixed-effects models that allowed inclusion of available data. Participants who dropped out also had a shorter duration of tinnitus and higher baseline anxiety symptoms, suggesting that attrition may not have been completely random. Second, program engagement was low and highly variable, with a substantial proportion of participants recording no usage. This limits conclusions about the effectiveness of Tune Out when used as intended and highlights the need for further investigation of adherence strategies. Third, the waitlist control condition controlled for the passage of time but did not control for non-specific intervention factors such as expectation, attention, or engagement with educational material. Additionally, participants were aware of their allocation, which may have influenced self-reported outcomes. Finally, the sample was self-selected and required internet access, which may limit generalisability to people with lower digital literacy, limited access to technology, or lower motivation to engage in online tinnitus support.

Additional limitations relate to the exploratory analyses. The regression, program usage, sensitivity, and self-efficacy analyses were not the primary focus of the trial and were likely underpowered. These findings should therefore be viewed as hypothesis-generating. Program usage was also self-selected, meaning that observed associations between engagement and outcomes cannot be interpreted causally.

The findings have several clinical and research implications. Unguided iCBT-based programs such as Tune Out may provide a useful low-intensity option within a stepped-care model of tinnitus management. Such programs could be offered to people awaiting specialist care, those who prefer self-guided support, or those who face barriers to accessing tinnitus-specific CBT. However, the engagement findings suggest that unguided access alone may not be sufficient for all users. Some individuals may benefit from additional guidance, reminders, or brief clinician-supported elements to help them initiate and sustain program use. Future research should examine whether guided, minimally guided, or adaptive versions of Tune Out improve adherence and outcomes compared with a fully unguided format.

Future studies should also include larger samples, longer follow-up periods, and active comparator conditions to better establish the effectiveness and durability of Tune Out. Research should examine mechanisms of change, including whether improvements are mediated by reduced tinnitus-related threat appraisal, increased sense of control, reduced arousal, or improved acceptance. Further work is also needed to identify who is most likely to benefit from unguided iCBT for tinnitus and what supports are needed for those less likely to engage.

## Conclusion

In conclusion, this study provides preliminary evidence that Tune Out, an unguided, self-paced online tinnitus management program, may reduce tinnitus severity and tinnitus handicap compared with waitlist control. The clearest effects were observed for tinnitus-related outcomes, while effects on broader psychological symptoms were less consistent. Usability ratings were favourable among respondents, but program engagement was highly variable and typical use was low. These findings support the potential value of unguided iCBT-based tinnitus programs as accessible low-intensity interventions, while highlighting the need for strategies to improve engagement and further research to confirm efficacy in larger and more diverse samples.

## Data Availability

All data produced in the present study are available upon reasonable request to the authors

